# Preterm birth and cardiometabolic health trajectories from birth to adulthood: the Avon Longitudinal Study of Parents and Children

**DOI:** 10.1101/2023.03.31.23287998

**Authors:** Gemma L Clayton, Laura D Howe, Linda M O’Keeffe, Adam J Lewandowski, Deborah A Lawlor, Abigail Fraser

## Abstract

**Background:** Adults who were born prematurely (<37 weeks gestation) are at increased cardiovascular disease risk, but it is unclear when in the life course this risk emerges. Our aim was to compare trajectories of multiple cardiometabolic risk factors from childhood to early adulthood between those who had and had not been born preterm.

**Methods:** Multilevel models were used to compare trajectories from early childhood (ranging from birth to 9 years) to age 25 years of BMI, fat mass, lean mass, systolic and diastolic blood pressure (BP), lipids, glucose and insulin, between participants born preterm (N=311-733, range 25-36 completed weeks gestation) and term (N=5365-12097) in a contemporary UK birth cohort study. We also investigated gestational age as a continuum.

**Results:** In children born preterm (versus term), systolic and diastolic BP were higher at age 7 (mean predicted differences 0.6mmHg; 95%CI -0.3, 1.5 and 0.6mmHg; 95%CI 0.03, 1.3, respectively). By age 25 years, the difference in SBP persisted (1.4, 95%CI -0.1, 2.9 mmHg) and in DBP (−0.2, 95%CI -1.3, 0.9 mmHg) disappeared. Participants born preterm (versus term) had lower BMI between 7 and 18 years, but by age 25, there was no difference. Lean mass and fat mass (measured from age 9 only) trajectories were consistent with BMI. HDL-c was higher, and triglycerides lower at birth in those born preterm, but this difference also disappeared by 25 years. There was no evidence of differences in glucose and insulin between participants born preterm compared to term.

**Conclusions:** There were few, modest differences in cardiometabolic health measures during early life in those born preterm versus term. All disappeared by age 25, except the small difference in SBP. Longer follow-up is needed to establish if and when trajectories of measures of cardiometabolic health in term and preterm born people diverge.

**Clinical perspective:** 

**What is new?:**

- Whether life course trajectories of commonly assessed cardiovascular disease (CVD) risk factors such as blood pressure, are different in people born preterm vs those born at term, is unknown.
- By age 25, we generally found no evidence of differences between people born preterm and term on measures of cardiometabolic health except that systolic blood pressure was modestly higher in those born preterm. We also observed more favourable outcomes with lower adiposity measures between ages 9 and 18 years.

**Clinical implications:**

- The reported increased risk of CVD in people born preterm is not apparent in early adulthood.
- Whilst it may emerge in later life, our results suggest no justification for CVD screening using ‘classic’ risk factors in young adults based on their gestational age.
- Further work to replicate these findings in other independent cohorts and studies with follow-up into mid life are required to examine when associations emerge.

## Introduction

Some 10% of babies worldwide are born pre-term (defined as <37 completed weeks of gestation) (1). Several national registry linkage studies have found that adults born preterm are at increased risk of cardiovascular disease (CVD) including stroke and myocardial infarction (MI) (2-5). Associations between preterm birth and adverse levels of CVD risk factors in early adulthood such as an increased risk of hypertension (6, 7), higher diastolic blood pressure (8, 9), higher lipid levels (4, 10, 11) and higher body mass index (BMI) (12-15) have also been reported. A recent systematic review reportied that adverse cardiometabolic consequences may not be limited to estreme and very preterm individuals (<28 and between 28 to 32 weeks respectively), with moderate to late preterm birth (between 32-36 weeks of gestation) also associated with increased risks of hypertension and diabetes (16). More adverse cardiac structure and function in people born preterm compared with terms have been reported (17, 18), although studies with more also tend to have more limited sample sizes.

Although there is consistent evidence that preterm birth is associated with CVD and its risk factors, it is less clear when associations emerge in the life course. looking at preterm birth and early life course trajectories of those risk factors would help better understand mechanisms and pathways that lead to increased CVD risk in later life, specifically what risk factors may be involved and when such associations emerge.

The aim of this study was to compare trajectories of change in multiple cardiometabolic risk factors from childhood to early adulthood between those who had and had not been born preterm in the early 1990s.. We also examine gestational age as a continuum. The cardiometabolic risk factors that we study are BMI, fat and lean mass, systolic blood pressure (SBP), diastolic blood pressure (DBP), pulse rate, triglycerides, high density lipoprotein cholesterol (HDL-c), non-HDL-c, glucose and insulin.

## Methods

### Study participants

Data from the Avon Longitudinal Study of Parents and Children (ALSPAC) were used. All pregnant women resident in the area surrounding the city of Bristol, United Kingdom (UK), who had an estimated delivery date between 1 April 1991 and 31 December 1992 were eligible for the study (19). Briefly, ALSPAC initially enrolled a cohort of 14,451 pregnancies, from which 13,867 live births occurred in 13,761 women. Follow-up has included parent and child completed questionnaires, links to routine data and clinic attendance. Research clinics were held when the participants were approximately seven, nine, 10, 11, 13, 15, 18 and 25 years old. Ethical approval for the study was obtained from the ALSPAC Ethics and Law Committee and the Local Research Ethics Committees. Full details of recruitment, follow-up and data collection for these women have been reported elsewhere (19, 20), and the study website contains details of all the data that is available through a fully searchable data dictionary and variable search tool (http://www.bristol.ac.uk/alspac/researchers/our-data/).

### Gestational age

Gestational age was determined from the last menstrual period but adjusted to reflect the early pregnancy ultrasound estimation if the two differed by two weeks or more, according to the clinical protocol in use at the time. Preterm birth was defined as delivery greater than or equal to 24 and before 37 completed weeks of gestation. Pregnancies with a recorded gestational age of greater than 41 weeks were excluded as there is evidence of worse fetal outcomes in prolonged gestations (21).

### Study outcomes

**Table 1** summarises the ages at which each risk factors was assessed and the sample size at each time point.

### Anthropometry

BMI (weight (kg) divided by height squared (m^2^)) was calculated from one to 25 years using data from several sources including research clinics, routine child health clinics, health visitor records and questionnaires. Central fat and lean mass were derived from whole body dual energy X-ray absorptiometry (DXA) scans using a Lunar prodigy narrow fan beam densitometer from 9 to 25 years (at ages nine, 11, 13, 15, 18 and 25).

### SBP, DBP and pulse rate

SBP, DBP and pulse rate were measured from 7 to 25 years (ages seven, nine, 10, 11, 13, 15, 18 and 25) at least twice each with the participant sitting and at rest with the arm supported, using a validated device and a cuff size appropriate for the upper arm circumference. The mean of the two final measures at each data collection timepoint was used here.

Blood based biomarkers

### Lipids

Total cholesterol, HDL-c and triglycerides were measured in cord blood at birth and from venous blood subsequently at ages seven, nine, 15, 18 and 25 years.. Non-HDL-c was calculated by subtracting HDL-c from total cholesterol at each measurement occasion. Samples were non-fasted at seven and nine; fasting measures were available from clinics at 15,18 and 25 years.

### Insulin and glucose

Insulin was measured on cord blood at birth. Non-fasting glucose was measured at age seven as part of metabolic trait profiling, using Nuclear Magnetic Resonance (NMR) spectroscopy. Fasting glucose and insulin were also available for a random 10% of the cohort at age nine years. Fasting glucose and insulin were measured at 15, 18 and 25 years.

### Confounders and other variables of interest

Confounders were defined *a priori using directed acyclic diagrams (22)*. We adjusted analyses for sex and maternal characteristics: age, self-reported pre-pregnancy BMI (kg/m^2^), smoking (any smoking during pregnancy vs not), parity (0, 1, 2, ≥3 pregnancies), alcohol intake during pregnancy (any/none), maternal education and hypertensive disorders of pregnancy (HDP).Maternal education was defined by the highest attained qualification (i) Certificate of Secondary Education (CSE), ordinary-(O-) level or vocational certificate (qualifications usually obtained at age 16, the UK minimum school leaving age when these women were at school), (ii) Advanced A-level (usually taken at 18 years) or (iii) university degree. Information on pre-pregnancy BMI, number of previous pregnancies and maternal education were obtained by self-report around the time of recruitment (during pregnancy at around 12 weeks gestation) to the study (mean age 28.3, SD 4.8). Maternal smoking status and alcohol intake before/during pregnancy was also self reported in pregnancy questionnaires up to 30 weeks gestation. As there were few (up to 56) cases of gestational diabetes (GDM) these women were excluded.

We performed additional sensitivity analyses adjusting for birthweight to explore whether we were primarily capturing differences in preterm birth or differences in birthweight. Birthweight was recorded by ALPSAC research staff after delivery or abstracted from medical records. However, given the intrinsic link between gestational age and birthweight whether the latter is a confounder or a mediator is debatable..

### Missing data

Missing values of maternal confounders were imputed due to ∼ 35% (4477/12830) missingness (**Table S1**). These were imputed using multivariable multiple imputation with chained equations, performed using the mi impute command in Stata 16. Those with complete data were more likely to have a higher education, more likely to be older and less likely to smoke during pregnancy or drink alcohol (see **Table S1**). We used 50 imputed data sets (with 25 iterations) and included all variables in the imputation models along with a number of additional auxiliary variables (predictive of missingness) (see **Table S2**).

The amount of missing data and the characteristics before and after imputation are presented in **Table S3**.

### Statistical analysis

We used multilevel models to examine and compare trajectories of cardiometabolic health of participants born preterm to those born at term, and by gestational age as a continuous variable. Multilevel models estimate mean trajectories of the outcome while accounting for non-independence of measures within individuals and include all participants with at least one cardiometabolic measure, under the missing at random (MAR) assumption.

Trajectories of cardiometabolic health up to age 18 by sex were modelled (23), and described in detail, (24) previously. Here, we extended these existing models (23) to include data from the 25-year clinic. Further details of how these trajectories were extended are included in supplementary material (Supplementary information on multilevel models, **Tables S4** and **S5**). All trajectories except BMI were estimated using linear spline multilevel models and trajectories of BMI were modelled using fractional polynomial multilevel models. Briefly, linear splines allow knot points to be fit at different ages to derive periods in which change is approximately linear whilst fractional polynomials can allow for more complex relationships and involve raising age to many combinations of powers. Models were fit with an interaction term between age and an indicator for preterm (vs full term) or gestational age, adjusting for sex and maternal confounders as detailed earlier. Sex and maternal confounders were added into the model as main effects which adjusts for the association between the covariate and the outcome at baseline. Values of cardiometabolic risk factors that had a skewed distribution (BMI, fat mass, insulin, and triglyceride) were (natural) log transformed prior to analysis. We report these associations both as an absolue association (on the log scale) and the relative measure as a percentage (difference on the log scale, multiplied by 100). Fat mass and lean mass were adjusted for height using the time- and sex-varying power of height that best resulted in a height-invariant measure to ensure at all ages lean and fat mass are independent of height (23, 25). All trajectories were modelled in MLwiN version 3.04, called from Stata version 16 using the runmlwin command. Model fit was assessed by comparing the mean observed values to the mean predicted values for each outcome by preterm/full term across age. We report the adjusted estimates for maternal age, smoking, alcohol, BMI, parity, sex, education and HDP from the multiple imputation datasets as the main results.

To ascertain whether or not the associations between gestational age (continuous) and each outcome were linear within each spline period, we compared models using a likelihood ratio test with gestational age split into quartiles and included as a single variable with models in which quartiles of gestational age was included as three dummy variables. These terms were fit as interactions with age within each spline period. We conducted a number of sensitivity analyses. We report both unadjusted and adjusted results following multiple imputation in **Supplementary material**. We also report a complete case analysis. We included births >41 gestational weeks (n up to 934) to evaluate potential selection bias arising from excluding these participants from our main analysis. We also ran a model in which we additionally adjusted for birth weight to investigate whether any associations between gestational age and outcomes was driven by size at birth.

## Results

Maternal characteristics by preterm birth (range 25-36 weeks gestation) and term (range 37-41 weeks gestation are shown in **Table 2**. Individuals born preterm were of a lower birth weight than individuals born at term. Mothers of preterm and term babies were similar ages (27.8 vs 28.1 years); but a higher proportion of mothers to preterm babies were first time mothers and had lower levels of education compared to mothers of term babies. HDP was more common amongst mother who delivered preterm (24.6 vs 16.0%).

There was no evidence for a departure from linearity in the association between gestational age (continuous) and all outcomes (**Table S6**). **Figures 1 and 2** show associations of preterm birth and gestational age with cardiovascular risk factors respectively, adjusted for all measured confounders (coefficients are presented in **Table 3** and **Tables S7-10**). **Figure 3** presents the associations of preterm birth adjusted for birthweight. Results from other models (sensitivity analyses) are presented in **Tables S11-21**. Observed and predicted values across each spline period and first and last time points were very similar for all outcomes (**Tables S22-25**).

**Figure 1.**
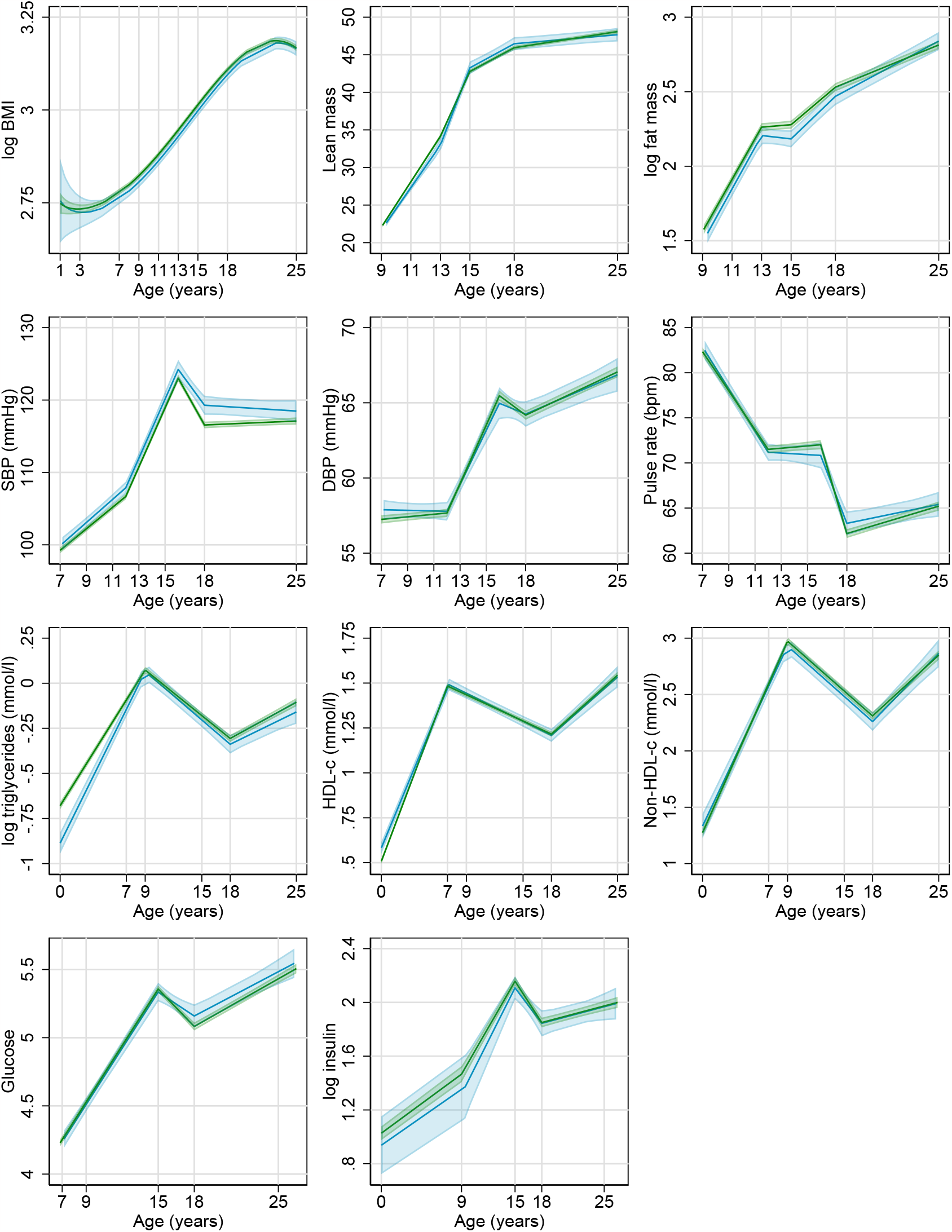
A Predicted mean (95% CI) trajectories of cardiometabolic risk factors by preterm and B full term birth and B Estimated mean difference (95% CI) between those born preterm versus full term. Full terms restricted to less than 42 weeks. All models adjusted for offspring sex and maternal characteristics: age, parity, smoking status, alcohol intake, pre pregnancy BMI and HDP. Trajectories drawn for females, mean maternal age and pre pregnancy BMI, no HDP, no previous children, no maternal smoking or alcohol and educated to CSE/ Vocational/ O-level. Estimates <0 mean lower values for preterm vs term, MD=mean difference.

**Figure 2.**
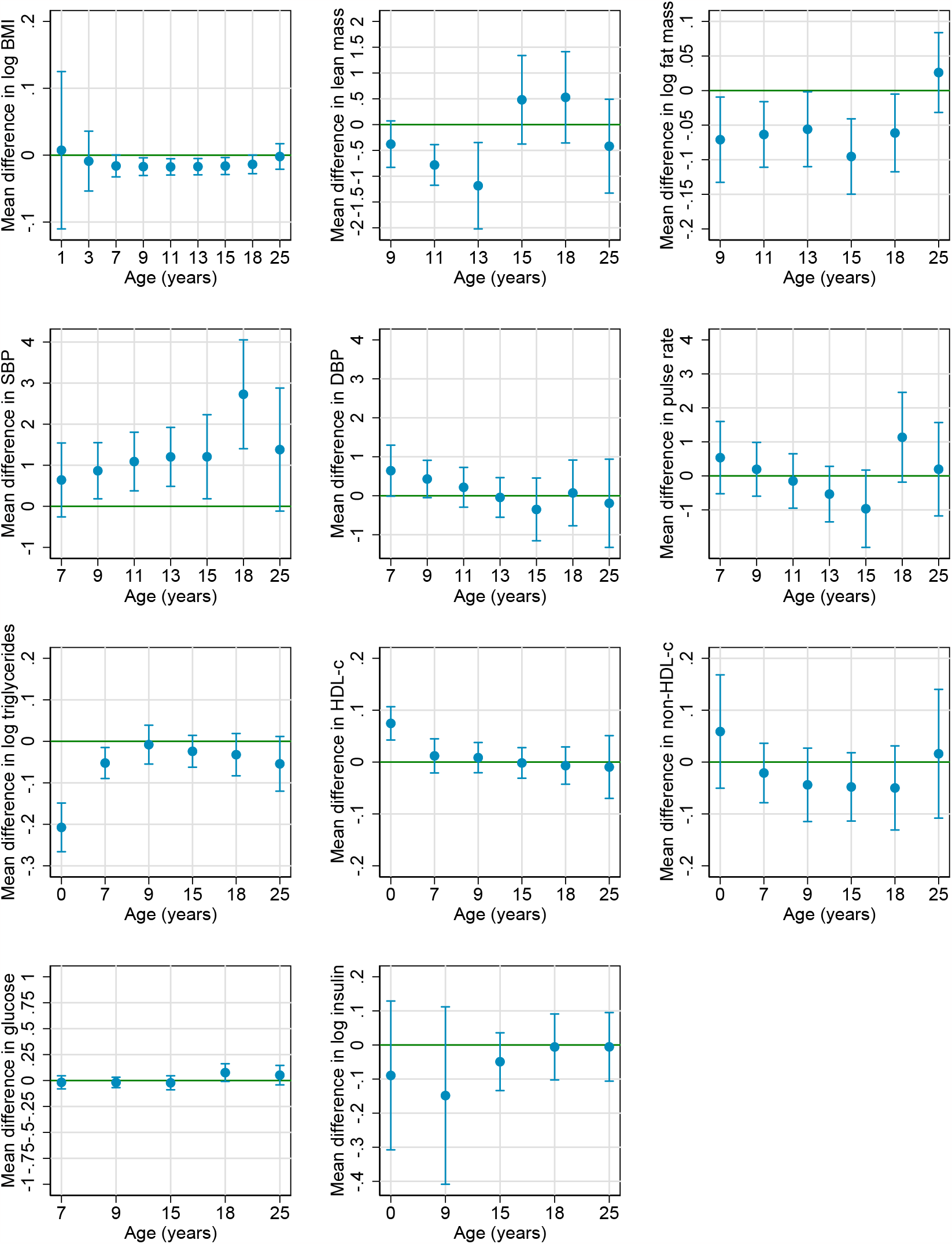
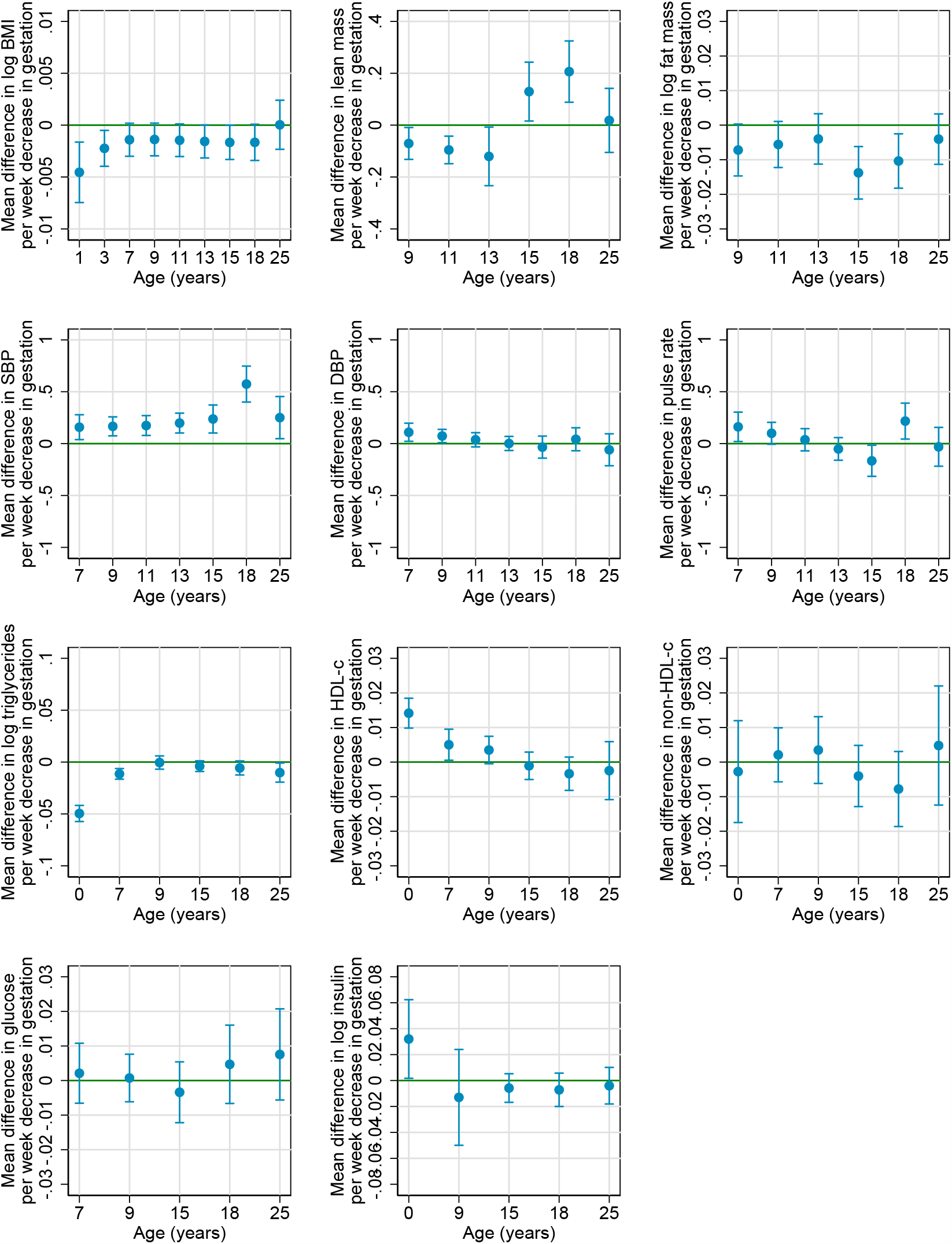
Estimated mean difference in cardiometabolic risk factors per 1 week decrease in gestational age. Gestational age restricted to less than 42 weeks. All models adjusted for offspring sex and maternal characteristics: age, parity, smoking status, alcohol intake, pre pregnancy BMI and HDP.

**Figure 3.**
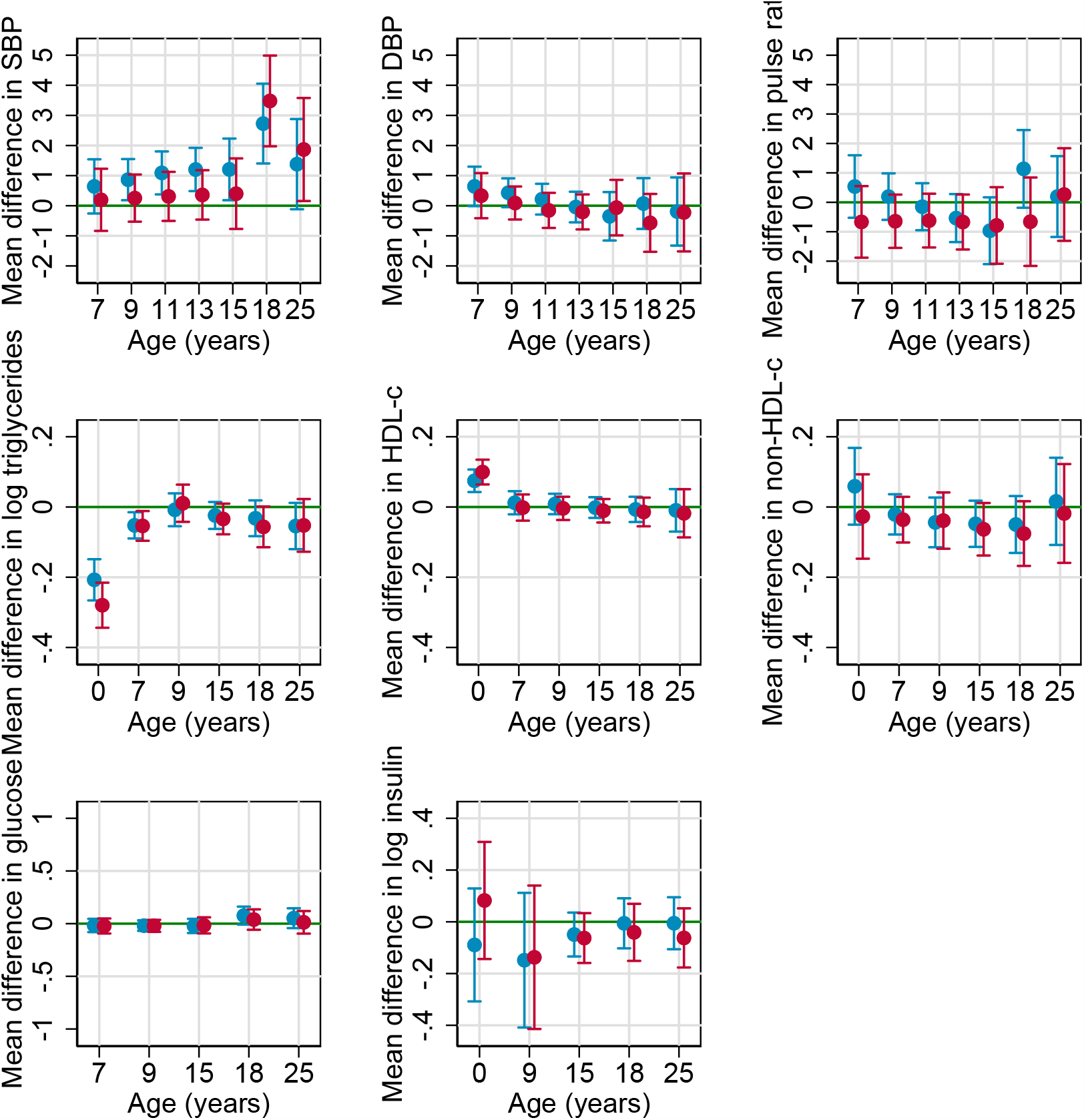
Estimated mean difference (95% CI) between those born preterm versus full term (blue) and additionally adjusted for birthweight (red) Full terms restricted to less than 42 weeks. All models adjusted for offspring sex and maternal characteristics: age, parity, smoking status, alcohol intake, pre pregnancy BMI and HDP. Estimates <0 mean lower values for preterm vs term, MD=mean difference.

Overall, the underlying shape of BMI from birth to early adulthood was J shaped, decreasing from age one to seven, then increasing until age 18 before it begins to plateau by age 25 (**Figure 1A and Table S11**). Predicted mean BMI was similar in participants born preterm compared to participants born at term at ages 1, 3 and 25 whilst participants born preterm had a lower mean BMI between the ages of 7 and 18 years (**Figure 1B**). (Height-adjusted) lean and fat mass (measured from age 9 only) trajectories were consistent with BMI. For example, mean fat mass was 7% (95% confidence interval (CI), (2,12) lower in preterm participants compared to those born at term, at age 15. Results were consistent when gestational age was modelled continuously (**Figure 2**) and when analyses included pregnancies of 42 weeks or more (**Tables S11-S13**).

SBP and DBP generally increased from age 7 to 16, before decreasing until age 18 and then beginning to plateau (**Figure 1A**). Both SBP and DBP were higher at age 7 (mean predicted differences 0.6mmHg; 95%CI, -0.3, 1.5 and 0.6mmHg; 95%CI, 0.03, 1.3, respectively) in preterm compared to term participants (**Figure 1B and Table 3**). From age 16 to 18 years, SBP decreased in both groups but at a slower rate in preterm participants, meaning that by age 18 mean SBP was 2.7mmHg higher (95% CI, 1.4, 4.1) in preterm participants (**Table 3**). The difference in SBP persisted (1.4, 95%CI, -0.1, 2.9 mmHg) and in DBP (−0.2, 95%CI, -1.3, 0.9 mmHg) attenuated to the null by 25 years. When modelling gestational age as a continuum (**Figure 2, Table S14**) these differences were also apparent, for example, mean SBP was 0.21 mmHg (95%CI, 0.03, 0.40) higher at age 25 per 1 week decrease in gestational age.

HDL-c was higher, and triglycerides lower at birth in those born preterm, but this difference persisted for triglycerides and attenuated to the null for HDL-c by 25 years (**Figure 1B**). Glucose appeared to be slightly higher at ages 18 and 25 years (**Figure 1B**). No other differences were found (**Figures 1 and 2, Table 3**). Furthermore, results were consistent when gestational was modelled continuously (**Figure 2**).

In sensitivity analyses, when pregnancies >41 weeks were included (**Tables S11-21**), results were consistent. Results were also similar in the complete case and multiple imputation analyses (**Tables S11-21**). When adjusting for birthweight, results remained similar across all risk factors (**Figure 3**). For example, results attenuated at later ages for glucose suggesting birthweight could be an explanation whilst they remained for SBP suggesting preterm birth is likely driving this association.

## Discussion

### Main findings

In this paper, we examined longitudinal changes in 11 measures of cardiometabolic health from early childhood or birth through to 25 years in participants born preterm compared to those born at term in a large contemporary birth cohort study. Some differences during early life were noted, namely lower height-adjusted lean and fat mass at age 9 and marginally higher SBP throughout from age 7 to 25 years.. Most of these differences attenuated to the null by age 25, but a small difference in mean SBP (1-2 mmHghigher) was observed in participants born preterm compared to term.

Given that we know that preterm born individuals have a higher risk of CVD (3, 7), several explanations for our results are possible. It is possible that in early life the cardiovascular risk factors measured here do not capture an already increased cardiovascular risk in people born preterm, and are better reflected using imaging modalities, such as retinopathy, which have been shown to differ in adolescents by gestational age (9). These differences may be reflected in divergent trajectories of BP and other cardiometabolic risk factors measured here after 25 years and in later adulthood (up to 37 years), and further follow-up of this and similar cohorts will therefore be valuable.

HDP are a leading cause of preterm birth (26, 27). Here, we were able to account for HDP (by adjusting for it as a potential confounder) and found that a small differences in offspring SBP persisted even when doing so. However, studies have shown that genetic variants associated with maternal BP are also associated with a shorter duration of gestation and preterm birth (27). This could mean that the association we find here between preterm birth and higher SBP are driven by genetic confounding rather than by exposure to preterm birth itself. Similarly, recent evidence from sibling and MR studies (28, 29) show that it is likely not the direct in utero exposure with HDP that affects the offspring cardiometabolic health but the maternal genes.

We also found evidence of favourable (lower) adiposity measures between ages 9 and 18 years in people born preterm. Whilst initially counter intuitive to higher BP, this inverse relationship of lower birth weight and higher later blood pressure has been attributed to genetic effects (30).

### Comparison to other studies

To the best of our knowledge, no other study has examined trajectories of cardiometabolic risk factors over time in adults born preterm versus term or by gestational age. Our findings suggest that those born preterm have a lower lean and fat mass at 9 years compared to full terms and is in line with other studies (31-33). For example, Fewtrell et al. measured body composition using DXA in 497 children born preterm (gestational age at birth: 25–36 weeks), compared them to 95 term-born controls at the age of 8.8–12.7 years (mean age: 11.2 years) and showed fat mass was lower in preterms.

Our findings are also broadly consistent (although we see a much weaker association) with a population-based cohort study of individuals born in 1985–1989 in Northern Finland (34) which found that preterm birth is associated with higher SBP at mean age 23 (SD=1.4) in a study of 134 preterm participants (3.2mmHg 95%CI (1.1, 5.4) and 242 late preterms (1.5mmHg (−0.3, 3.3) compared to 334 term born participants. A meta-analysis by Parkinson et al, included 17,030 preterm and 295,261 term-born adults from twenty seven studies, also found higher blood pressure in those born preterm compared to full term (32). Bertagnolli et al suggests that the higher SBP in those born preterm (compared to full term) arises from greater vascular stiffness in adults born preterm (12). Elastin synthesis in arterial walls occurs at the end of gestation and a shorter gestation can disrupt this process and cause elastin disruption which in turn increases vascular stiffness leading to an increase in SBP and ultimately greater risk of hypertension. The meta-analysis by Parkinson et al also showed no evidence of an association in insulin or glucose between those born pre-terms compared to full term (32).

### Strengths and limitations

Strengths of this study include the availability of repeat measures of multiple cardiometabolic risk factors over a 25 year period in a large contemporary, population based cohort – and the ability to look at change over time. We also used multiple imputation to account for missing confounder data and included participants with at least one measurement in the analysis under the MAR assumption, so that selection bias was minimised. In our main analysis we compared participants born preterm (<37 weeks) to term (37-41 weeks). However, in sensitivity analyses we excluded up to 934 women with a gestational age >41 weeks and our results remained very similar. We also note that participants in ALSPAC are predominantly of White European origin, and previous studies have shown ethnic differences in cardiometabolic risk factors, so our findings might not be generalisable to other race/ethnic groups.

Despite the size of the cohort, we were unable to stratify preterm birth further into very/extremely preterm due to the small number of participants. However, we did examine gestational age as a continuous variable. We checked for a departure from a linear association between gestational age in weeks and the cardiometabolic health outcomes and did not find strong evidence of this. Our results across all outcomes were still apparent and estimated more precisely compared to when gestational age was categorised as preterm/full term. However, we acknowledge this may not help if extreme preterm are very different and given the small numbers, it is unlikely we would find statistical evidence of departure from linearity even if it is real. Given that previous studies have shown sex differences in cardiometabolic risk factors it is possible that associations with preterm birth also differ by sex. However, given the relatively small number of people born preterm (140-330 in females and 171-403 in males) we were unable to explore sex differences.

Given that low birth weight is most often caused by being born preterm it is possible that rather than observing associations of preterm birth we are also observing associations between low birth weight and cardiovascular health. It is difficult to unpick gestational age from gestational size due to them being so closely related and that it is likely that cause and effect differ amongst those born preterm (35, 36). However, including birthweight in the main model made little to no difference to the gestational age/preterm associations.

## Conclusion

The known increased risk of CVD seen in adults born preterm was not apparent based on classic CVD risk factors in our early adulthood cohort except for a modest increase in systolic blood pressure. We also observed more favourable outcomes with lower adiposity measures between ages 9 and 18 years. Reducing preterm birth would be unlikely to have substantial impact on improving conventional cardiometabolic risk factors during the first 25 years of life. Further work to replicate these findings in other independent cohorts and studies with follow-up into mid life are required to examine when associations emerge.

## Supporting information

Main tables

## Data Availability

Access to ALSPAC data is through a system of managed open access (http://www.bristol.ac.uk/alspac/researchers/access/).

## Availability of data and materials

Access to ALSPAC data is through a system of managed open access

(http://www.bristol.ac.uk/alspac/researchers/access/).

Analysis scripts can be found on the following GitHub page:

https://github.com/gc13313/CVDinpretermoffspring.

## Supplementary information on multilevel models

Individual-level random effects allow the intercepts and slopes for each period to differ between individuals and therefore each model was fit with random effects at each knot point. For lean mass, due to few available repeated measures, we modelled the person-specific random effects as a single linear slope rather than a function of the splines as was done in all other linear spline models. This allowed person specific variation from the average trajectory but under the assumption that person-specific deviation from the mean trajectory was constant over time. An age and height adjusted covariate was included as a fixed effect in fat/lean mass models.

As we also included data from the 25-year clinic we checked whether an additional knot point was needed at 18 years for all risk factors except BMI. We compared models using model fit statistics including Akaike Information Criterion (AIC), Bayesian Information Criterion (BIC) and the likelihood ratio test to select the best fitting model (Tables S4 and S5).

BMI has been modelled previously using fractional polynomials and is described elsewhere (1). With the addition of the 25 year data we used fractional polynomials again to help select the best fitting curve Briefly, BMI was log transformed due to skewness of the data and fractional polynomials were used where age was raised to various combinations of powers (each of the following single powers, plus each combination of two powers: 0.5, 1, 2, 3, -0.5, -1, -2, natural log), from which we selected the best fitting curve (the one with the lowest likelihood value). The resulting curve contained three age terms including log age, log age* age and log age *age^2.

